# Lysosomal polygenic risk score in Parkinson’s disease across populations

**DOI:** 10.64898/2026.09.14.26362981

**Authors:** Wenhua Sun, Kathrin Brockmann, Isabel Wurster, Roswitha Kemmner, Benjamin Röben, Reda Skaiene, Stefanie Lerche, Claudia Schulte, Thomas Gasser, Manuela Tan, the Global Parkinson’s Genetic Program (GP2)

## Abstract

**Background:** Lysosomal dysfunction has been implicated in Parkinson’s disease (PD) pathogenesis. However, lysosomal polygenic risk scores (lyso-PRS) have not been evaluated extensively in PD across populations.

**Objectives:** (1) Evaluation of lyso-PRS (lyso-PRS) performance for PD risk across populations; (2) Comparison of lyso-PRS across idiopathic PD (iPD), *GBA1*-PD, non-manifesting *GBA1* carriers (NMC), and healthy controls (HC) in the European population. (3) Assessment of the lyso-PRS association with clinical outcomes in the PD patients of European population.

**Methods:** We analyzed Neurobooster array data from the Global Parkinson’s Genetics Program (GP2) dataset, including 32,054 iPD cases and 18,188 healthy controls across ten ancestries.

**Results:** (1) lyso-PRS performed best in Europeans (European-GP2 test cohort: AUC = 0.602, p = 2.02e-172; European-TUEPAC validation cohort: AUC = 0.632, p = 5.34e-20) and worse in other populations (AUC: 0.51-0.59, p<0.05). (2) In the European population, the lyso-PRS distinguished *GBA1*-PD from HC (AUC = 0.621, p = 7.83e-71) and from NMC (AUC = 0.621, p = 1.05e-18). (3) In the European-GP2 cohort, higher lyso-PRS was significantly associated with earlier age at onset (β=-0.21, p=0.022) and higher Unified Parkinson’s Disease Rating Scale (UPDRS) scores (β=0.32-1.13, all p < 0.05).

**Conclusions:** lyso-PRS was associated with iPD and *GBA1*-PD status, with the strongest predictive performance in the European population, underscoring the need to improve accuracy in other ancestries. In the European population, the lyso-PRS showed associations with clinical features; however, not all findings were replicated in the validation cohort, highlighting the need for further investigation in large and diverse cohorts.

## Introduction

Parkinson’s disease (PD) is clinically heterogeneous, with core motor features of bradykinesia, rest tremor and rigidity, accompanied by a wide range of non-motor symptoms (e.g., cognitive decline, sleep disturbance, depression, and automatic dysfunction). ^1^ The etiology of PD is complex, arising from an interplay of genetic factors, aging and environmental exposures that promote the a-synuclein misfolding and aggregation, ultimately leading to neurodegeneration. ^2^

The latest genome-wide association study (GWAS) uncovered numerous common genetic risk variants associated with PD, collectively explaining approximately 20% of heritability of PD in the European population. ^3^ While each single GWAS hit can only confer a small proportion of disease susceptibility, polygenic risk scores (PRS) integrate the cumulative effects of many variants based on GWAS results. ^4^ Given that more than 90% of cases are idiopathic PD (iPD), PRS represents a useful approach for characterizing the polygenic architecture in iPD and further identifying individuals with higher genetic risk of developing PD. Applying PRS to a special gene set that regulates biological functions known to be altered in PD could potentially help elucidate the contribution of specific molecular pathways to disease susceptibility and their impact in determining the clinical phenotype.

Pathway-specific PRS extends general PRS by restricting analysis to biologically defined gene sets, thereby enabling assessment of the cumulative effect of common risk alleles within specific molecular pathways implicated in PD. This approach may offer mechanistic insight beyond genome-wide PRS by linking inherited susceptibility to specific biological processes. Indeed, mitochondrial pathway PRS has recently been associated with increased PD risk in both European and Ashkenazi Jewish ancestries, ^5–7^ supporting the utility of pathway-specific PRS models.

Lysosomal dysfunction has been implicated in PD pathogenesis. ^8^ Converging evidence indicates impaired lysosomal function, including structural abnormalities of lysosomes, reduced immunoreactivity of lysosome-associated proteins, and accumulation of undegraded autophagosomes in postmortem brain tissue. ^9, 10^ *GBA1* is a major lysosomal risk locus and variants in *GBA1* are now recognized as a common strong genetic risk factor for PD. Deficiency in the activity of the enzyme glucocerebrosidase (Gcase), encoded by *GBA1*, leads to accumulation of ceramides within lysosomes, contributing to lysosomal dysfunction and impaired protein degradation. ^11, 12^

Genetic studies indicate that neurodegeneration in PD cannot be attributed solely to aging and environmental factors, underscoring a broad contribution of genes linked to lysosomal function in PD. Previous work has shown a significant excessive burden of rare, likely damaging variants in genes associated with lysosomal storage disorders among PD patients without known pathogenic variants, independent of *GBA1*. ^13^ In addition, lysosomal polygenic risk scores (lyso-PRS) have been reported to be associated with increased PD risk ^14^ and faster progression of cognitive decline ^15^ in European PD patients after exclusion of the *GBA1* locus. Together, these findings provide a strong rationale for investigating lysosomal PRS as a pathway-specific measure of genetic susceptibility in PD.

We constructed a lyso-PRS and evaluated its performance in the Global Parkinson’s Genetics Program (GP2) release 11 dataset, comprising 32,054 iPD cases and 18,188 healthy controls across ten ancestries. We then examined the discriminatory ability of the lyso-PRS model in the European population by comparing healthy controls, non-manifesting carriers (NMC) and iPD against *GBA1*-PD. Moreover, we assessed the associations of lyso-PRS with clinical outcomes in a European-GP2 exploratory cohort and an independent European Tuebingen Parkinson validation cohort (European-TUEPAC). Together, these analyses aimed to identify the contribution of lysosomal polygenic burden to PD risk and clinical phenotype, and to allow patient stratification for precision medicine in lysosomal centered clinical trials.

## Methods

### Subjects

The Global Parkinson’s Genetics Program (GP2) (Global Parkinson’s Genetics, 2021) dataset contains 32,054 cases and 18,188 controls in ten ancestries: European [European-GP2: 21,328 cases, 9,574 controls; Tuebingen Parkinson cohort (European-TUEPAC): 2,122 cases, 451 controls], African American (AAC: 266 cases, 595 controls), African (AFR: 1,145 cases, 2,580 controls), Ashkenazi Jewish (AJ: 1,169 cases, 220 controls), Latino and indigenous Americas (AMR: 1,754 cases, 1,363 controls), Central Asian (CAS: 507 cases, 676 controls), East Asian (EAS: 2,486 cases, 1,448 controls), Middle Eastern (MDE: 580 cases, 681 controls), South Asian (SAS: 212 cases, 304 controls), Complex Admixture History (CAH: 485 cases, 296 controls). Table S1 summarizes the demographics of these eleven sub-cohorts.

In European ancestry individuals, there were 23,449 idiopathic PD (iPD), 2,206 *GBA1*-PD, 482 non-manifesting *GBA1* carriers (NMC) and 10,025 healthy controls (HC). Populations with less than 100 individuals (i.e. Finnish, 57 cases and 10 controls) were excluded. Cases and controls with missing covariates (including sex, age and first five principal components) were removed.

Participants’ clinical information and genetic samples were obtained with written consent and appropriate institutional ethical approvals.

### Genetic data

We used genetic data from the GP2 release 11 dataset(https://zenodo.org/records/17753486). This includes imputed genotype data generated using the Neurobooster array, ^16^ with the quality control conducted with Genools (https://github.com/GP2code/GenoTools) ^17^ at a sample and variant level, as detailed in the Supplementary Information 1.

Idiopathic Parkinson’s disease (iPD) was defined by excluding individuals carrying known pathogenic variants in established PD genes, including *GBA1*, *LRRK2*, *SNCA*, *VPS35*, *PARK7* (*DJ-1*), *PINK1*, *PRKN*, *RAB32*, *ATP13A2*, *DCTN1*, *DNAJC6*, *FBXO7*, *JAM2*, *RAB39B*, *SLC20A2*, *SYNJ1*, *VPS13C*, and *WDR45*. Carrier status for *GBA1* and *LRRK2* was determined by clinical exome sequencing (CES) and whole-genome sequencing (WGS) as well as genome-wide Illumina NeuroBooster Array genotyping data within the GP2 consortium within a large multicentric collaboration.^18^ For the remaining PD genes, pathogenic variant carriers were identified directly from the GP2 Illumina NeuroBooster Array genotype data. Healthy controls were also restricted to individuals without pathogenic variants in these genes. Data processing and quality control are described elsewhere. ^17^ For individuals with both sequencing and array data available, concordance checks were performed using CES/WGS calls to validate Neurobooster array genotyping results, particularly for rare variants. Pathogenic/likely pathogenic variants according to ClinVar and/or ACMG criteria are referred to as causative variants, and selected variants with an established PD-risk association are referred to as risk variants. Cases and controls carrying established *GBA1* risk-associated variants were classified as *GBA1*-PD and non-manifesting *GBA1* carriers (NMC), respectively.

### Clinical data

#### (1) European-GP2

Non-motor experiences of daily living, motor experiences of daily living, the severity of motor symptoms and motor complications were assessed using part I–IV of the Unified Parkinson’s Disease Rating Scale (UPDRS),^19^ respectively. Because GP2 participants were assessed across multiple studies, different instruments were used to evaluate the same symptom domains. Cognitive impairment was defined as a Montreal Cognitive Assessment (MoCA) ^20^ score < 26, Mini–Mental State Examination (MMSE) ^21^ score < 24, and scales for outcomes in Parkinson’s disease-cognition (SCOPA-COG) ^22^ score < 24. Depression was tested using Geriatric Depression Scale-15 (GDS-15) ^23^ and the Beck Depression Inventory (BDI). ^24^ REM Sleep Behaviour Disorder (RBD) was screened using the RBD Screening Questionnaire (RBDSQ). ^25^ Excessive daytime sleepiness was measured using the Epworth Sleepiness Scale (ESS). ^26^

#### (2) European-TUEPAC

UPDRS I-IV assessments were done as described in European-GP2. Cognitive function was assessed using MoCA ^20^ and MMSE scales. ^21^ The same MMSE and MoCA cutoff used in European-GP2 was applied to the European-TUEPAC cohort. Depressive symptoms were assessed using the Beck Depression Inventory II (BDI-II). ^24^

##### Lysosomal polygenic risk score

The polygenic risk score (PRS) was calculated as the weighted sum of the number of risk alleles carried by each individual. Allele weights were derived from 2025 PD genome-wide association study (GWAS) summary statistics in the European population. ^3^

Lysosomal polygenic risk scores (lyso-PRS) were calculated using PRSice-2 v2.3.5 (https://github.com/choishingwan/PRSice). ^27, 28^ Lysosomal genes were defined from the Molecular Signatures Database (MSigDB, https://www.gsea-msigdb.org/gsea/msigdb, RRID:SCR_016863) by selecting all 44 gene sets containing the term “lysosom*”, yielding 519 unique genes. Variants from the 2025 PD GWAS summary statistics were annotated to genes using ANNOVAR. ^29^ Gene mapping was restricted to variants located within the annotated genomic coordinates of each gene. Of the 519 lysosomal genes, 491 overlapped with genes represented in the GWAS summary statistics. Given the influence of *GBA1* and *LRRK2* on lysosomal biology and its close interactions with other lysosomal related genes, these two genes were excluded from the overlapping gene set.

Variants with minor allele frequency (MAF) <0.01 were excluded. After filtering, 217,053 SNPs were retained for lyso-PRS construction. Linkage disequilibrium (LD) and clumping was performed using default settings (r2 = 0.1 and distance of 250kb). The percentages of lysosomal SNPs by ancestry were detailed in Table S2. In the European ancestry individuals, the dataset was divided into a training set (European-GP2) and an independent testing set (European-TUEPAC). P-value threshold optimization was performed in the training set, and the optimal threshold (p = 0.1) was then applied to the testing set. In other ancestries, owing to limited sample size, PRSice-2 was employed using its default clumping-and-thresholding framework across four p value thresholds (P ≤ 0.001, 0.05, 0.2, 0.4). lyso-PRS were standardized within each ancestry before analysis.

Statistical analyses, including regression models, covariate adjustment, and meta-analysis, are detailed in Supplementary information 1. The overall study workflow is illustrated in Figure 1.

**Figure 1.**
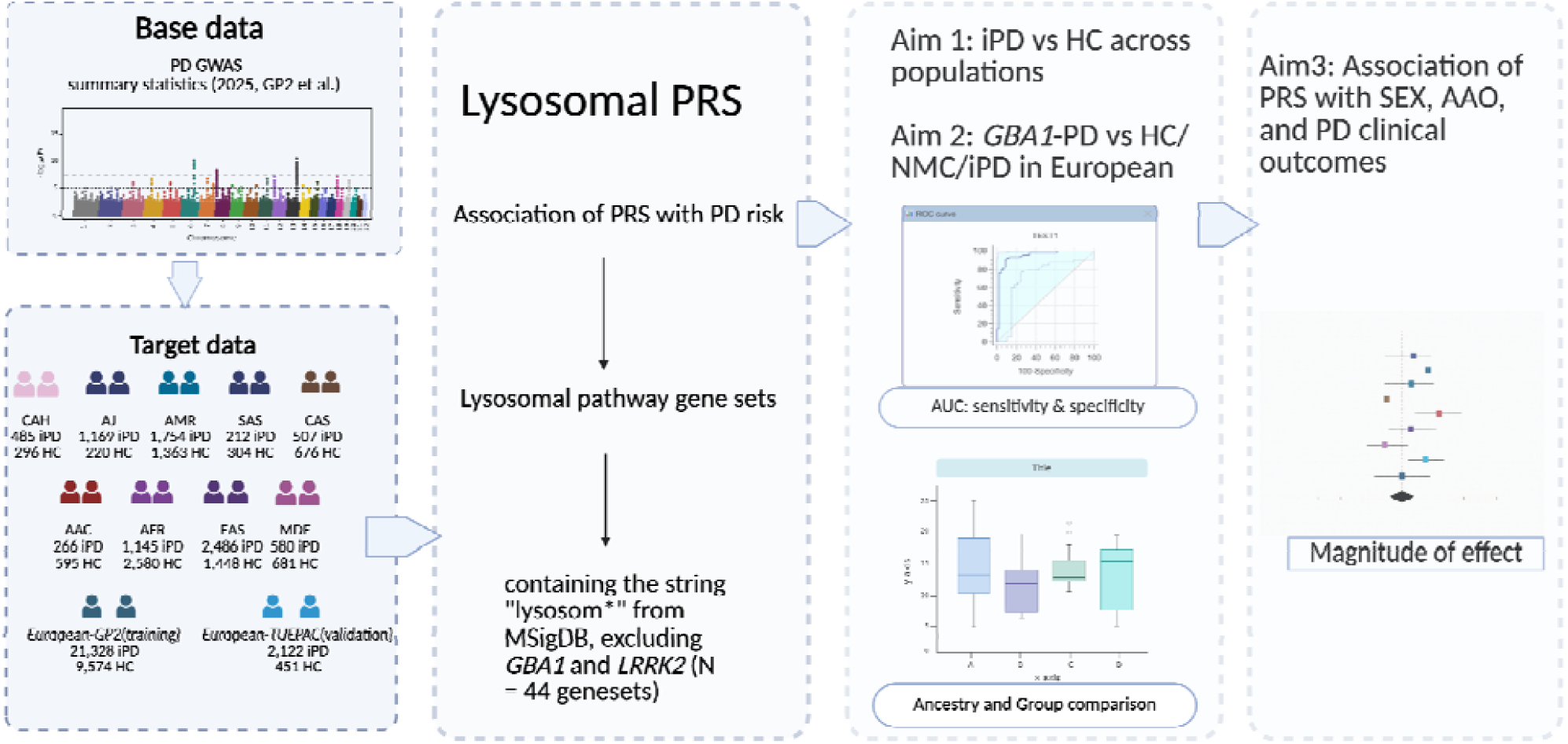
Workflow of analysis. Abbreviations: European-GP2, European ancestry NeuroBooster array cohort; European-TUEPAC, European ancestry Tuebingen Parkinson cohort; AJ, Ashkenazi Jewish; AMR, Latino and indigenous Americas; SAS, South Asian; CAS, Central Asian; AAC, African American; AFR, African; EAS, East Asian; CAH, Complex Admixture History; MDE, Middle Eastern; iPD, idiopathic Parkinson’s disease; HC, healthy controls; NMC, non-manifesting GBA1 carriers; AUC, the area under curve; AAO, age at onset.

## Results

### 1. Lysosomal polygenic risk score (lyso-PRS) performance across ancestries

The latest European GWAS ^3^ derived lyso-PRS models exhibited ancestry-dependent predictive performance for discriminating idiopathic PD (iPD) from healthy controls (HC) across ancestries, adjusted by sex, age and the first five genetic principal components (PCs) (Figure 2 and Table 1). Compared with other ancestries, predictive performance was highest in the European population (Figure 2). Specifically, the training cohort (Neurobooster array, European-GP2) achieved an area under the curve (AUC) of 0.602, OR of 1.45 (95% CI: 1.42-1.49), and balanced accuracy of 0.576, while the validation dataset (Tuebingen Parkinson’s cohort, European-TUEPAC) showed a higher AUC of 0.632, OR of 1.70 (95% CI: 1.52-1.90) and balanced accuracy of 0.589 (Table 1). In contrast, all non-European ancestries exhibited a reduced performance with AUCs below 0.6 (Table 1). The lowest performance of the lyso-PRS model was observed in the African ancestry (AUC: 0.509, OR (95% CI: 1.07 (0.99-1.15), balanced accuracy: 0.512) (Table 1).

**Figure 2.**
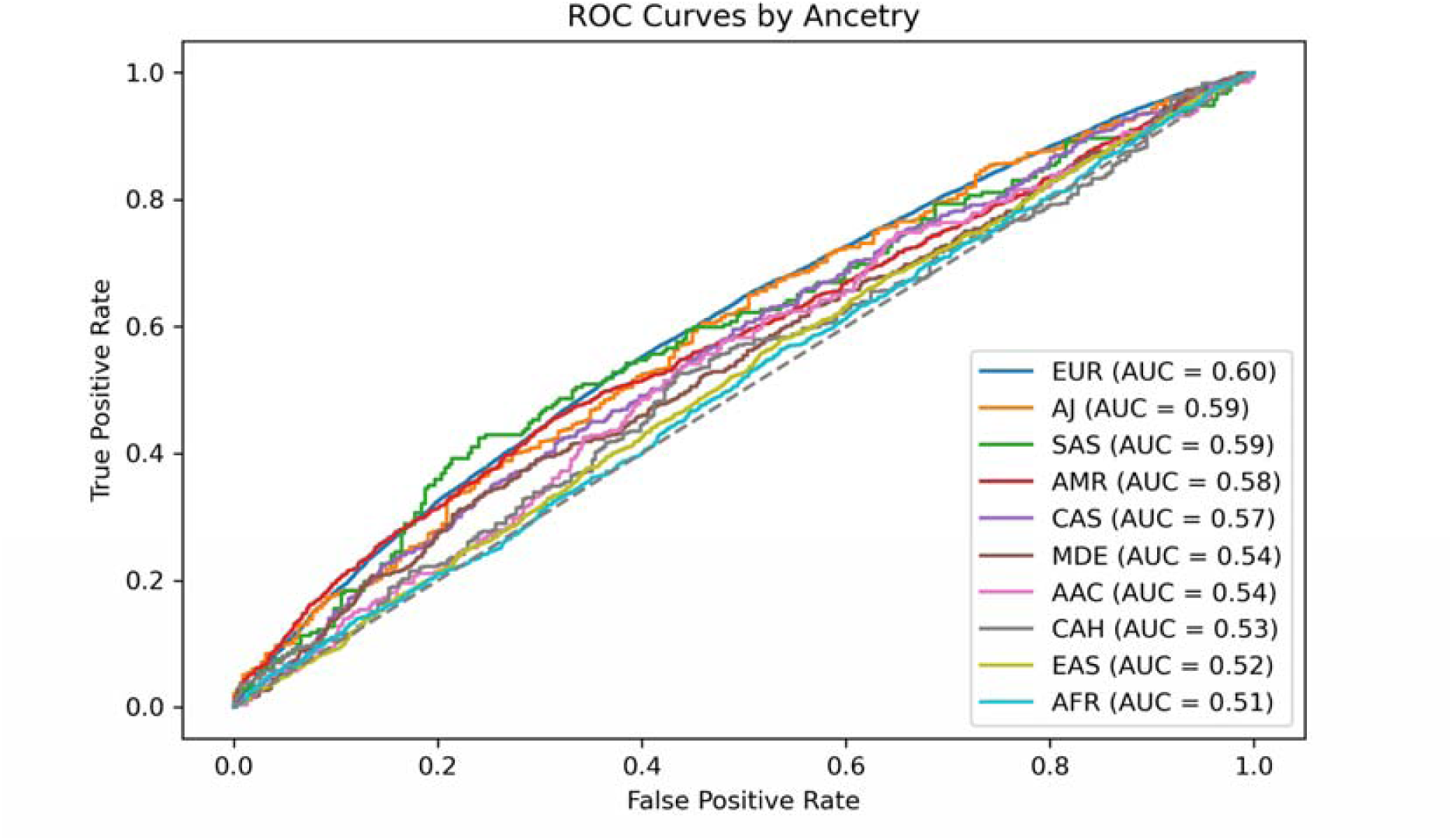
lyso-PRS performance for each ancestry. Receiver operating characteristic (ROC) curves evaluating the performance of lysosomal polygenic risk score (lyso-PRS).

**Table 1.**
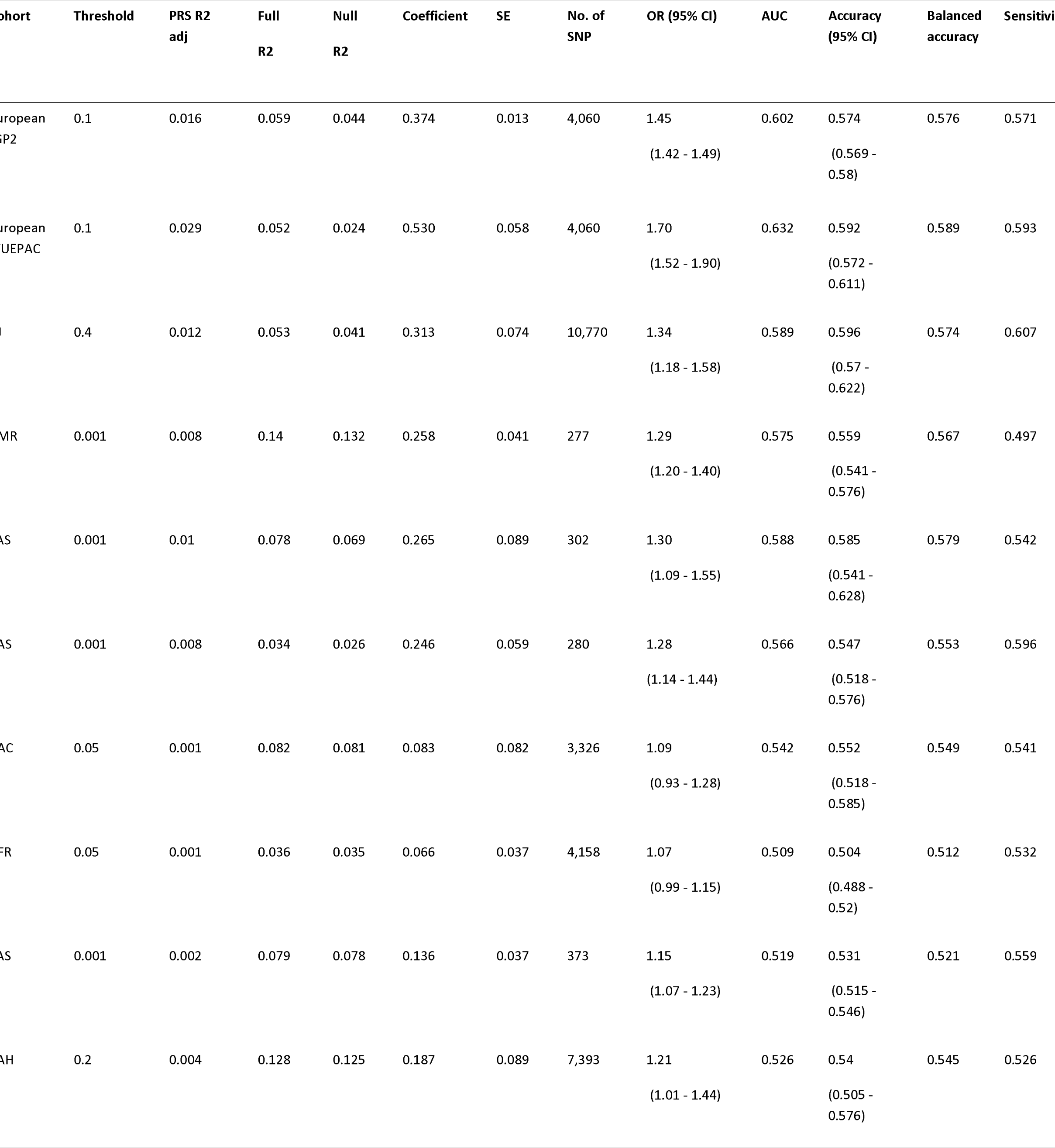
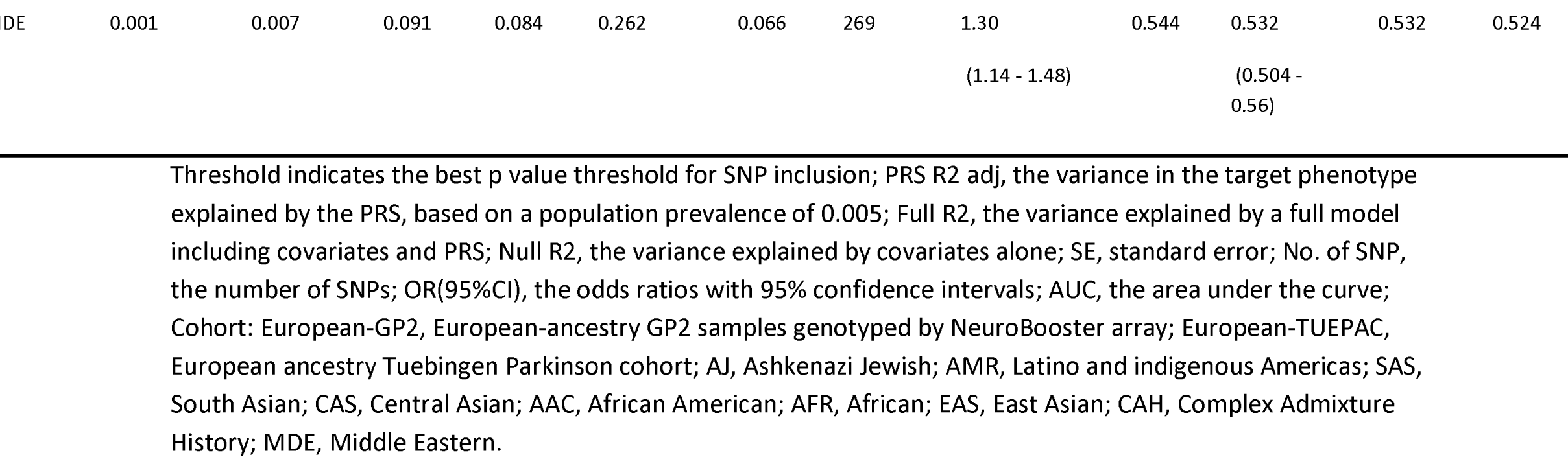
Summary statistics of lyso-PRS performance between iPD and controls across ancestries.

Further regression analyses were conducted to compare lyso-PRS between iPD and controls across ten ancestries. In the European-GP2 and European-TUEPAC groups, lyso-PRS scores were significantly higher in iPD patients than in healthy controls (European-GP2: Beta = 0.37, SE = 0.013, p = 2.02e -172, European-TUEPAC: Beta = 0.53, SE = 0.058, p = 5.34e -20). Similarly, in the most non-European groups, iPD patients had higher lyso-PRS scores compared to healthy controls (p < 0.05). However, no significant difference in lyso-PRS scores was observed between iPD and controls in the African American (Beta = 0.083, SE = 0.082, p = 0.31) and African ancestries (Beta = 0.066, SE = 0.037, p = 0.075) (Table S3).

### 2. lyso-PRS performance in the European population across *GBA1*-PD comparisons

In European ancestry individuals, lyso-PRS showed moderate performance in distinguishing *GBA1*-PD from healthy controls and non-manifesting carriers, with AUCs of 0.621 for both comparisons and balanced accuracies of 0.586 and 0.593, respectively. The estimates were OR of 1.56 for *GBA1*-PD versus HC and OR of 1.57 for *GBA1*-PD versus NMC. In contrast, the lyso-PRS model showed a weak performance between *GBA1*-PD and iPD, with an AUC of 0.515, an OR of 1.05, and a balanced accuracy of 0.513 (Figure 3A and Table S4). Consistent with these findings, logistic regression showed significantly higher lyso-PRS scores in *GBA1*-PD group than in NMC (Beta = 0.45, SE = 0.051, p = 2.05e-18) and in HC (Beta = 0.44, SE = 0.025, p = 7.83e-71) groups, but only a small difference relative to iPD (Beta = 0.049, SE = 0.022, p = 0.026) (Figure 3B and Table S3).

**Figure 3.**
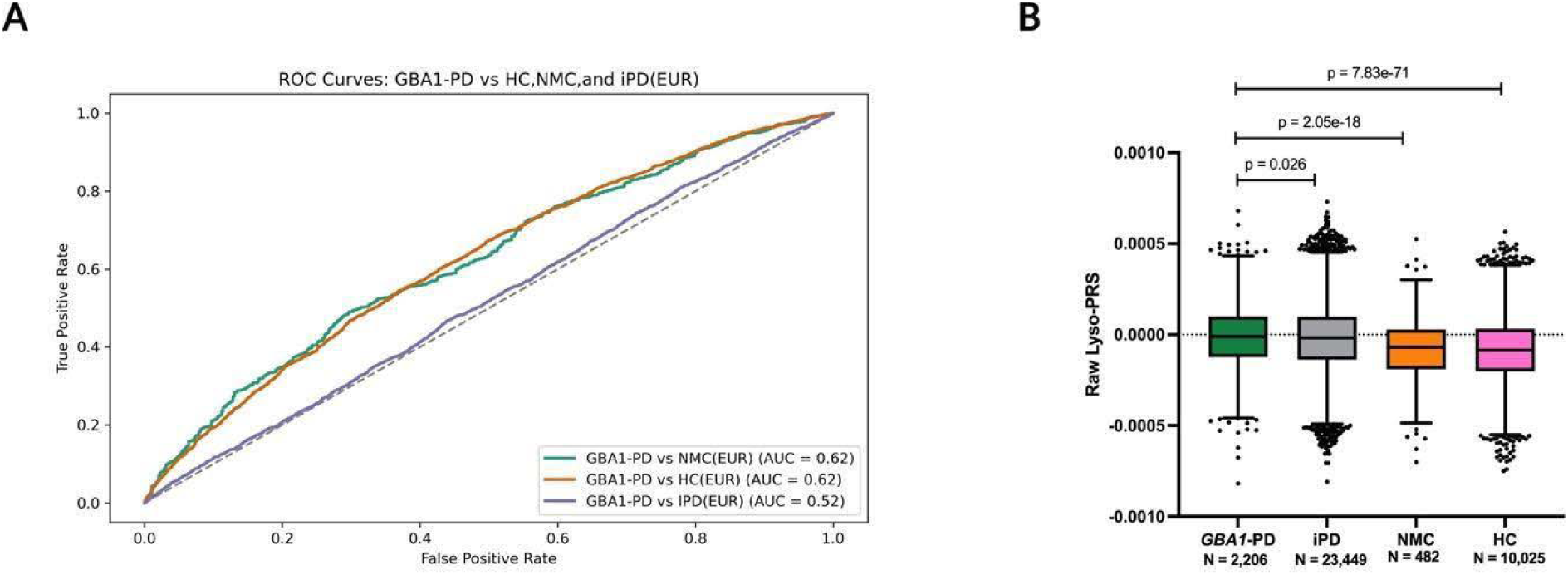
lyso-PRS performance in the European population across GBA1-PD comparisons with HC, NMC, and iPD. (A) Receiver operating characteristic (ROC) curves showing discrimination of *GBA1*-PD from non-manifesting GBA1 carriers (NMC), healthy controls (HC), and idiopathic Parkinson’s disease (iPD). (B) Box-and-whisker plots of raw lysosomal polygenic risk score (lyso-PRS) distributions, using the optimal P-value threshold (P_T = 0.1), across *GBA1*-PD, iPD, NMC, and HC groups in the European population.

### 3. Associations of lyso-PRS with sex and age at onset in iPD patients of European-GP2 (training) and European-TUEPAC (validation) cohorts

Given the multicenter composition of the European-GP2 cohort, which included participants from diverse regions, we first assessed the association of lyso-PRS with sex and age at onset in European studies. No significant association between lyso-PRS and sex was observed in either the training cohort, European-GP2 (23 studies, N = 20,162, Beta = 0.0094, SE = 0.016, random-effects p = 0.59), or the validation cohort, European-TUEPAC (N = 2,122, Beta = 0.064, SE = 0.043, p = 0.14) (Figure 4A, Table S5-S6).

**Figure 4.**
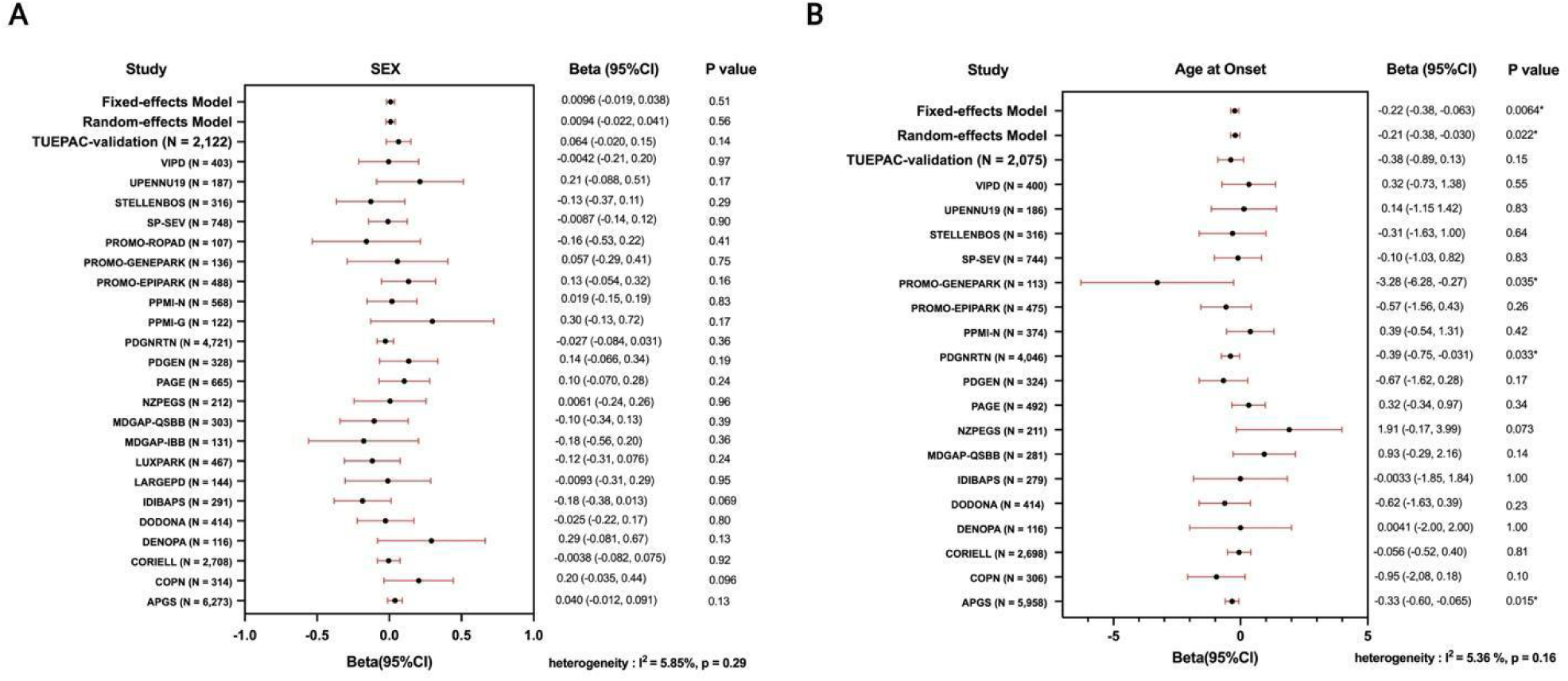
Forest plots summarize regression estimates with 95% confidence intervals for the lyso-PRS association with sex and age at onset across European-GP2 training cohort and European-TUEPAC validation cohort. (A) Meta-analysis of the association with sex across 23 sub-cohorts from the European-GP2 cohort and the European-TUEPAC cohort; (B) Meta-analysis of the association with age at onset across 18 sub-cohorts from the European-GP2 cohort and the European-TUEPAC cohort. Fixed-effect and random-effects summary estimates were shown for the European-GP2 multi-cohort analysis. European-GP2 referred to GP2 samples in the European population genotyped using the Illumina NeuroBooster Array; European-TUEPAC referred to European-ancestry samples from the Tübingen Parkinson dataset genotyped using the Illumina NeuroBooster Array.

Higher lyso-PRS was significantly associated with earlier age at onset after adjusting for covariates in the European-GP2 cohort (18 studies, N = 17,733, Beta = -0.21, SE = 0.090, random-effects p = 0.022). Although this association was not replicated in the European-TUEPAC cohort (N = 2,075, Beta = -0.38, SE = 0.26, p = 0.14), the direction of effect was consistent with the European-GP2 finding. (Figure 4B, Table S5-S6).

### 4. Associations of lyso-PRS with clinical outcomes in iPD patients of European-GP2 (training) and European-TUEPAC (validation) cohorts

Significant associations with UPDRS-I, UPDRS-II and UPDRS-III were observed only in the European-GP2 cohort (UPDRS-I: 3 studies, N = 1,100, Beta = 0.48, SE = 0.18, random-effects p = 0.0073; UPDRS-II: 3 studies, N = 1,105, Beta = 0.87, SE = 0.23, random-effects p = 1.34e-04; UPDRS-III: 5 studies, N = 1,847, Beta = 1.13, SE = 0.50, random-effects p = 0.024) and were not replicated in European-TUEPAC (UPDRS-I: N = 591, Beta = -0.30, SE = 0.25, p = 0.23; UPDRS-II: N = 579, Beta = -0.25, SE = 0.34, p = 0.47; UPDRS-III: N = 1,670, Beta = 0.25, SE = 0.36, p = 0.45) (Figure 5A-5C, Table S5-S6).

**Figure 5.**
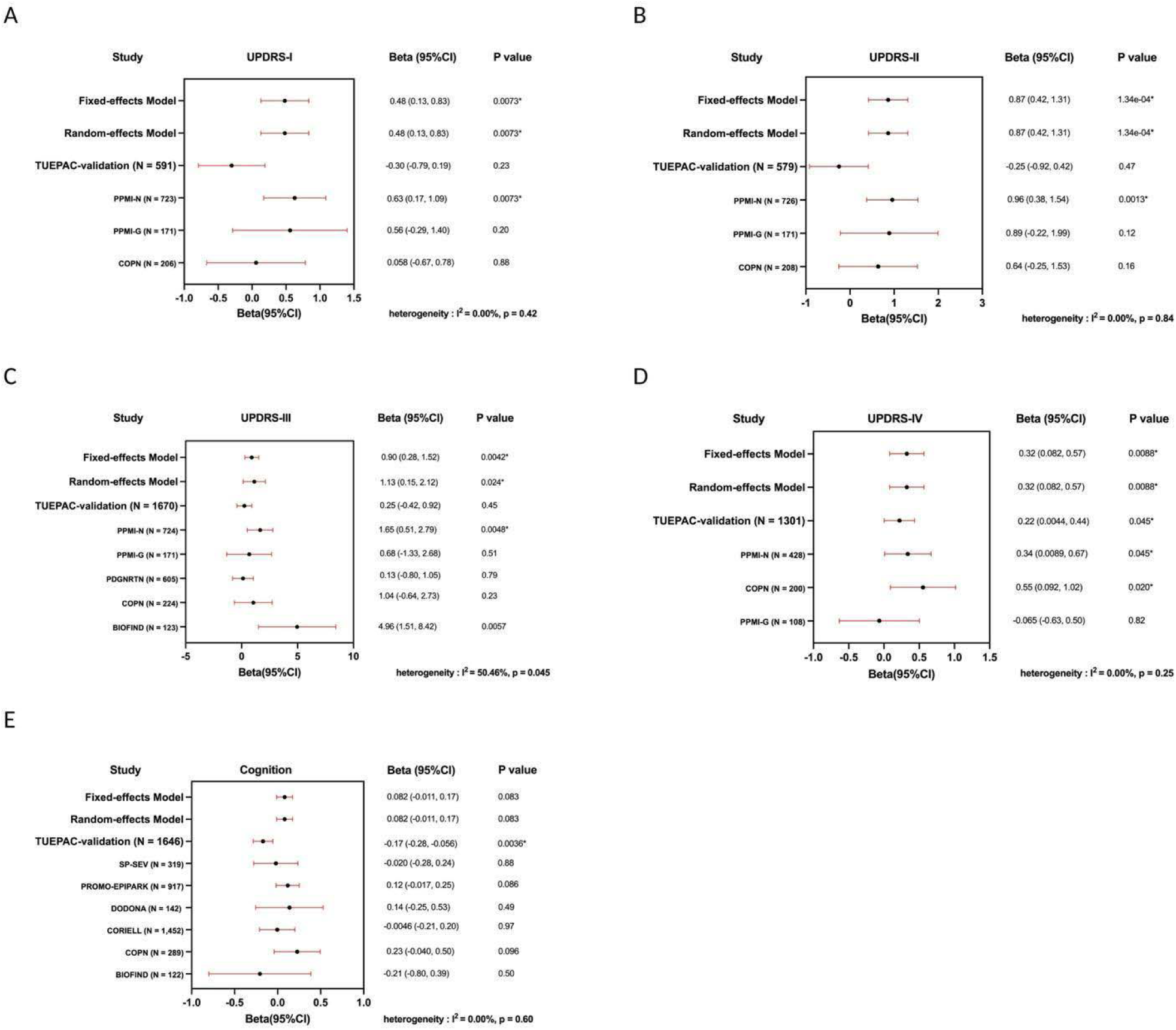
Forest plots summarize regression estimates with 95% confidence intervals for the association between lyso-PRS and clinical outcomes across European-GP2 training cohort and European-TUEPAC validation cohort. (A-D) UPDRS I-IV; (E) Cognition. Regarding cognitive impairment analyses, PD patients were classified as normal cognition or impaired cognition based on established cutoffs for the Mini-Mental State Examination (MMSE, <24), Montreal Cognitive Assessment (MoCA, <26), and Scales for Outcomes in Parkinson’s Disease-Cognition (SCOPA-COG, <24). European-GP2, GP2 samples in the European population genotyped using the Illumina NeuroBooster Array; European-TUEPAC, European ancestry Tuebingen Parkinson cohort; UPDRS, Unified Parkinson’s Disease Rating Scale.

In contrast, higher lyso-PRS was consistently associated with higher UPDRS-IV score in both the European-GP2 (3 studies, N = 736, Beta = 0.32, SE = 0.12, random-effects p = 0.0088) and European-TUEPAC cohorts (N = 1,301, Beta = 0.22, SE = 0.11, p = 0.045) (Figure 5D, Table S5-S6).

Regarding cognitive performance, no significant association with lyso-PRS was found in the European-GP2 training cohort (6 studies, N = 3,241, Beta = 0.082, SE = 0.047, random-effects p = 0.083). However, in the European-TUEPAC validation cohort, higher lyso-PRS was associated with worse cognition after adjustment for covariates (N = 1,646, Beta = -0.17, SE = 0.058, p = 3.58e-03) (Figure 5E, Table S5-S6).

Associations between lyso-PRS and non-motor symptoms, as well as sensitivity analyses additionally adjusting for disease duration, are detailed in Supplementary Information 1.

## Discussion

This study represents the largest and the first comprehensive evaluation of lyso-PRS for predicting PD risk and disease severity across ten populations. We found that lyso-PRS showed the strongest predictive performance in European ancestry compared to other ancestries. This predictive ability was further replicated in the independent European-TUEPAC validation cohort. Previous studies of lyso-PRS mostly focused on European populations.

Building on this work, our study provides a landscape of the contribution of lysosomal genetic burden to PD across populations. Interestingly, lyso-PRS distinguished PD case-control status independent of *GBA1* mutation status, suggesting that common lysosomal pathway variation may capture additional genetic risk beyond damaging *GBA1* variants. Finally, clinical outcomes regression analysis revealed higher lyso-PRS scores may be associated with worse performance of UPDRS scales and cognition decline in PD patients, although not all associations were replicated in the validation cohort.

Consistent with one previous finding, ^14^ we replicated lyso-PRS association with iPD risk in the large European Neurobooster array cohort comprising 21,328 patients and 9,574 controls. However, the predictive performance of the lyso-PRS was modest, with an AUC of approximately 0.60. Reduced and more variable predictive performance was observed in the other non-European populations. This pattern was likely influenced by the use of European summary statistics across all ancestry groups, as currently available non-European summary statistics remain underpowered to estimate robust effect sizes. ^30–32^ This highlighted the importance of robust population-specific summary statistics. Given the greater genetic diversity and differences in linkage disequilibrium patterns across populations, larger GWAS in non-European populations are needed to generate reliable summary statistics and improve the accuracy of polygenic risk prediction, particularly in underrepresented populations.

This study further examined the relevance of lyso-PRS in *GBA1*-associated PD, given the central role of glucocerebrosidase in lysosomal biology and its established contribution to PD pathogenesis. Notably, lyso-PRS also distinguished *GBA1*-PD from unaffected *GBA1* carriers, supporting that *GBA1* mutations alone are insufficient for disease manifestation, as also demonstrated by the reduced penetrance of these variants, and that the polygenic background may contribute to penetrance. This was consistent with prior work showing that polygenic background has been found to modulate the PD risk of *GBA1* variants in carriers of p.E365K, p.T408M and p.N409S variants. ^33^

By contrast, lyso-PRS did not differ between *GBA1*-PD and iPD, with a low AUC of 0.51. Together with the comparisons involving *GBA1*-PD, we found lyso-PRS was more strongly associated with PD status instead of *GBA1* mutation status. Previous work revealed *GBA1* mutations and PRS independently increase PD risk, with PRS exerting an additive effect on the penetrance of *GBA1* mutations. ^34^ Taken together, our findings raise the possibility that *GBA1* mutations and common lysosomal polygenic burden contributed additively to PD susceptibility. Therefore, a deep understanding of the relationship between *GBA1* mutation and lysosomal polygenic background in PD can be expected to be helpful in the future to integrate lyso-PRS and *GBA1* into PD genetics for better risk stratification.

To clarify how lysosomal polygenic burden shaped the clinical phenotype of PD, we examined associations between lyso-PRS and a range of clinical outcomes. In a meta-analysis of 23 independent studies comprising 20,162 PD patients, we found no significant association between lyso-PRS and sex, and this null finding was also observed in European-TUEPAC. In addition, lyso-PRS was associated with earlier age at onset in a meta-analysis of 18 studies including 17,733 PD patients, which was not replicated in the European-TUEPAC. This discrepancy may reflect limited statistical power due to the smaller sample size of the European-TUEPAC cohort, as well as potential regional heterogeneity.

Regarding motor features, higher lyso-PRS was associated with worse motor experiences of daily living measured by UPSRS-II and motor severity assessed by UPDRS-III in the European-GP2 cohort, although this finding was not replicated in the European-TUEPAC. Previous studies have largely reported negative results for associations between lyso-PRS and UPDRS-II and UPDRS-III, including studies with 305 cases ^14^ and approximately 3,000–5,000 cases, ^35^ respectively. The partial inconsistency between the European-GP2 and European-TUEPAC cohorts, as well as with prior reports, may reflect limited statistical power. Only a limited number of cohorts were available for meta-analysis of most clinical outcomes, which may have reduced statistical power and contributed to instability of effect estimates. In addition, methodological differences across studies, including the selection of lysosome-related variants and PRS construction parameters, may also have influenced the results.

For non-motor features, higher lyso-PRS was associated with worse non-motor experiences of daily living measured by UPDRS-I in the European-GP2 cohort, but this association was not confirmed in the European-TUEPAC cohort. In contrast, higher lysosomal polygenic burden was associated with poorer cognitive performance in the European-TUEPAC cohort, consistent with prior work, ^15^ although this finding was not statistically significant in the European-GP2 cohort. Several factors may account for this discrepancy. The European-TUEPAC cohort is relatively homogeneous, whereas the multicenter European-GP2 cohort includes PD patients from diverse regions. Additionally, the use of different cognitive assessment tools (SCOPA-COG, MMSE, and MoCA) across cohorts may contribute to the inconsistent results. Overall, our findings suggest that lysosomal genetic burden beyond *GBA1* may influence specific motor and non-motor manifestations of PD, although these effects were not uniformly observed across cohorts. More broadly, pathway-specific PRS may better capture molecular or pathological biomarkers than complex clinical outcomes, which are inherently heterogeneous and influenced by multiple biological and environmental factors.

Interestingly, lyso-PRS was positively associated with UPDRS-IV, reflecting the severity of motor complications, in both cohorts.To our knowledge, this is the first time to find that dopaminergic motor complications were related with lysosomal polygenic risk. Previous works have reported that motor complications were more common in patients with pathogenic *GBA1* variants, ^36, 37^ and *GBA1*-mediated lysosomal and lipid dysregulation has been proposed as a key pathogenic axis in PD. ^12^ Together, our findings extend these observations by suggesting that, beyond *GBA1* variants, a broader lysosomal genetic burden may also contribute to the development or severity of motor complications. These results support a potential link between lysosomal dysfunction and treatment-related motor complications in PD. Further studies are essential to validate this association and to clarify whether lyso-PRS could help identify patients at higher risk of developing motor complications. Therefore, lyso-PRS could serve as therapeutic targets to reduce the risk and delay the time of motor complications during dopaminergic treatment.

Notably, previous work has suggested a surprising overlap between the non-motor manifestations of PD and lysosomal storage disorders (LSDs). ^38^ In this context, our results provide further support for a role of lysosomal dysfunction in shaping selected clinical features of PD. Future advanced large-scale studies integrating pathway-specific genetic burden with molecular biomarkers and detailed clinical assessment of motor and non-motor symptoms will be essential to better define the clinical relevance of lysosomal dysfunction and to clarify the mechanisms linking lysosomal pathways to phenotypic heterogeneity in PD.

We acknowledge that our study has some limitations. Firstly, the scarcity of non-European summary statistics necessitated the use of European-based data to construct the lyso-PRS, which may have reduced transferability and introduced bias in non-European populations. Second, *GBA1* carrier classification was challenging because rare variants of *GBA1* gene were difficult to genotype accurately from Neurobooster array-based data, particularly given the presence of the highly homologous pseudogene *GBAP1*. Although available WGS and CES data were used for concordance checks, residual carrier misclassification cannot be excluded. In addition, despite the relatively large European dataset, the sample size remained limited for meta-analyses of clinical outcomes, thereby reducing power to detect effects. Finally, we could not get access to more clinical data in non-European populations, and this precluded a more comprehensive cross-ancestry assessment of these associations.

In conclusion, this study is, to our knowledge, the largest cross-population analysis of lysosomal polygenic risk in PD to date. Our findings extend current understanding of how lysosomal common genetic variants contribute to PD risk and clinical features. These results further support an important role for lysosomal biology in PD and highlight the potential utility of pathway-specific polygenic profiling for biological stratification. With additional validation and improved cross-ancestry data, lyso-PRS may help identify high-risk PD patients to benefit from therapeutic strategies targeting lysosomal dysfunction.

## Data and Code Availability Statement

Data used in the preparation of this article were obtained from the Global Parkinson’s Genetics Program (GP2; https://gp2.org). Specifically, we used Tier 2 data from GP2 release 11 (https://zenodo.org/records/17753486). GP2 data are available on AMP PD (https://amp-pd.org).

Data of European Tuebingen Parkinson cohort dataset are available in an anonymized format on request to.

All code generated for this article, and the identifiers for all software programs and packages used, are available on GitHub (https://github.com/GP2code/lyso-PRS) and were given a persistent identifier via Zenodo (10.5281/zenodo.22682252).

## Acknowledgements

This project was supported by the Global Parkinson’s Genetics Program (GP2; https://gp2.org), a research program of Aligning Science Across Parkinson’s (ASAP) (https://ror.org/03zj4c476). ASAP is managed by the Coalition for Aligning Science (CAS) (https://ror.org/00rf4ez40). The Michael J. Fox Foundation for Parkinson’s Research (MJFF) (https://ror.org/03arq3225) is the funding and strategic partner. For a complete list of GP2 members see https://doi.org/10.5281/zenodo.7904831. A full list of members and their affiliations also appears in the Supplementary Information 2. For the purpose of Open Access, the author has applied a CC BY public copyright licence to any Author Accepted Manuscript version arising from this submission.

W.S. acknowledges the support of the China Scholarship Council program (Project ID: 202207040033).

## Contributions

(1) Research project: A. Conception, B. Organization, C. Execution; (2) Statistical analysis: A. Design, B. Execution, C. Review and critique; (3) Manuscript preparation: A. Writing of the first draft, B. Review and critique.

W.S.: 1A, 1B, 1C, 2A, 2B, 2C, 3A, 3B

K.B.: 1C, 3B

I.W.: 1C, 3B

R.K.: 1C, 3B

B.R.: 1C, 3B

R.S.: 1C, 3B

S.L.: 1C, 3B

C.S.: 1A, 1B, 1C, 2C, 3B

T.G.: 1C, 3B

M.T.: 1A, 1B, 2A, 2C, 3B

GP2: data generation, curation, and resource provision.

