## Supplementary Information 1 for "Lysosomal polygenic risk score in Parkinson’s disease across populations"

### Methods

Quality control of Neurobooster array data

At the sample level, quality control included call rate outliers (--mind, 0.05), biologic sex mismatches (--check-sex, default cutoffs: 0.25 ≤ F ≤ 0.75), relatedness check (--grm-cutoff, in GCTA; Usually set at 0.125 to remove first cousins or more related individuals), and heterozygosity rate outliers (--het, default range: −0.15 ≤ F ≤ 0.15). At the variant level, quality control included missingness by case control (--test-missing, using P > 1e-04), missing by haplotype (--test-mishap, using P > 1e-04), Hardy–Weinberg equilibrium (--filter-controls --hwe, using P > 1e-04) and variant missingness (--geno, 0.05).

Prevalence estimates for *GBA1*-PD comparisons

For the *GBA1*-PD versus NMC analysis, the prevalence was defined as the estimated prevalence of *GBA1*-PD among *GBA1* variant carriers. Based on previously reported estimates ranging from 3% to 30%, we used a conservative prevalence estimate of 5%. For the *GBA1*-PD versus iPD analysis, prevalence was defined as the proportion of *GBA1*-PD among all PD cases. As this proportion has been reported to range from approximately 5% to 15%, we used an estimate of 10%. For the *GBA1*-PD versus HC analysis, prevalence was defined as the estimated prevalence of *GBA1*-PD in the general population. This was calculated as the overall PD prevalence multiplied by the estimated proportion of *GBA1*-PD among PD cases: 0.005 × 0.10 = 0.0005. Because p.T408M and p.E365K carriers were included among *GBA1* carriers, and these variants are more frequent but have lower penetrance, we applied an additional adjustment factor of 0.10. Therefore, the final prevalence estimate used for this analysis was 0.005 × 0.10 × 0.10 = 0.00005.

Statistical analysis

Analyses and visualization were performed using R v4.3.3 and GraphPad prism 10.2.0. Associations between lyso-PRS and disease status were evaluated using multivariable logistic regression adjusted for age, sex, and the first five genetic principal components (PCs). Predictive performance was assessed with the area under the receiver operating characteristic curve (AUC).

Associations between lyso-PRS and sex across the ten ancestries were assessed using multivariable logistic regression adjusted for the first five PCs. Associations between lyso-PRS and age at onset (AAO) were assessed using multivariable linear regression adjusted for sex and the first five PCs.

Because of limited sample size, analyses of associations between lyso-PRS and motor and non-motor clinical scales were restricted to the European-GP2 training cohort and the European-TUEPAC test cohort. Age at visit, sex, and the first 5 genetic PCs were used as covariates.

Results from different independent studies within the European-GP2 cohort were combined using random-effects meta-analysis with metafor version 4.8-0 package in R. Heterogeneity of effects were assessed using Cochran’s Q and I^2^ statistics.

All analyses were performed using R statistical language and Python programming language on the Verily workbench (<https://workbench.verily.com/>).

### Results

Regression analysis of non-motor symptoms

In addition, we did not find that any of the four additional clinical outcome measures (Depression related measures, GDS-15, RBDSQ and ESS) were significantly associated with lyso-PRS in either the European-GP2 or the European-TUEPAC cohorts (Figure S1A-S1D, Table S5-S6).

Sensitivity analyses

We additionally included disease duration as a covariate in sensitivity analyses (Table S7, S8 and Figure S2). Because disease duration was not available for all individuals, the sample size for UPDRS analyses was substantially reduced, potentially limiting statistical power. After adjustment for disease duration, the associations of lyso-PRS with UPDRS-I and UPDRS-III in the European-GP2 cohort were no longer significant (UPDRS-I: N = 573, Beta = 0.10, SE = 0.23, random-effects p = 0.67; UPDRS-III: N = 911, Beta = 0.40, SE = 0.59, random-effects p = 0.50). In the European-TUEPAC cohort, the association between lyso-PRS and UPDRS-IV was attenuated and did not remain significant after adjustment for disease duration (N = 1,301, Beta = 0.18, SE = 0.098, random-effects p = 0.070).

### Supplementary Figures and Tables

Table S1. Demographics of cohorts across ten populations and European-TUEPAC.

| Cohort |  | Controls | iPD |
| --- | --- | --- | --- |
| AAC | N | 595 | 266 |
|  | Male (%) | 214 (36%) | 148 (56%) |
|  | Mean age at genotype [SD] (years) | 63.7 [13.1] | 67.4 [11.8] |
|  | Mean age at onset [SD] (years) |  | 61.1 [12.3] |
| AFR | N | 2,580 | 1,145 |
|  | Male | 1,322 (51%) | 787 (69%) |
|  | Mean age at genotype [SD] (years) | 62.0 [12.4] | 66.0 [9.9] |
|  | Mean age at onset [SD] (years) |  | 60.5 [9.8] |
| AJ | N | 220 | 1,169 |
|  | Male | 115 (52%) | 794 (68%) |
|  | Mean age at genotype [SD] (years) | 67.0 [10.5] | 71.7 [9.0] |
|  | Mean age at onset [SD] (years) |  | 64.8 [10.5] |
| AMR | N | 1,363 | 1,754 |
|  | Male | 467 (34%) | 986 (56%) |
|  | Mean age at genotype [SD] (years) | 59.8 [8.8] | 63.1 [12.2] |
|  | Mean age at onset [SD] (years) |  | 55.0 [13.1] |
| CAH | N | 296 | 485 |
|  | Male | 122 (41%) | 282 (58%) |
|  | Mean age at genotype [SD] (years) | 47.8 [19.1] | 62.8 [12.8] |
|  | Mean age at onset [SD] (years) |  | 55.9 [13.9] |
| CAS | N | 676 | 507 |
|  | Male | 300 (44%) | 224 (44%) |
|  | Mean age at genotype [SD] (years) | 59.9 [9.8] | 62.0 [11.0] |
|  | Mean age at onset [SD] (years) |  | 54.0 [12.1] |
| EAS | N | 1,448 | 2,486 |
|  | Male | 677 (47%) | 1,320 (53%) |
|  | Mean age at genotype [SD] (years) | 62.5 [11.3] | 67.9 [10.0] |
|  | Mean age at onset [SD] (years) |  | 59.4 [11.6] |
| MDE | N | 681 | 580 |
|  | Male | 301 (44%) | 325 (56%) |
|  | Mean age at genotype [SD] (years) | 61.3 [13.3] | 65.0 [10.9] |
|  | Mean age at onset [SD] (years) |  | 55.7 [12.6] |
| SAS | N | 304 | 212 |
|  | Male | 196 (64%) | 136 (64%) |
|  | Mean age at genotype [SD] (years) | 56.3 [13.7] | 64.1 [11.4] |
|  | Mean age at onset [SD] (years) |  | 56.0 [12.9] |
| European-GP2 | N | 9,574 | 21,328 |
|  | Male | 4,442 (46%) | 13,493 (63%) |
|  | Mean age at genotype [SD] (years) | 62.5 [13.7] | 68.5 [10.2] |
|  | Mean age at onset [SD] (years) |  | 60.8 [11.5] |
| European-TUEPAC | N | 451 | 2,122 |
|  | Male | 219 (49%) | 1,359 (64%) |
|  | Mean age at genotype [SD] (years) | 67.7 [6.9] | 65.0 [10.8] |
|  | Mean age at onset [SD] (years) |  | 58.6 [11.3] |

European-GP2, GP2 samples in the European population genotyped using the Illumina NeuroBooster Array; European-TUEPAC, European ancestry Tuebingen Parkinson cohort; AJ, Ashkenazi Jewish; AMR, Latino and indigenous Americas; SAS, South Asian; CAS, Central Asian; AAC, African American; AFR, African; EAS, East Asian; CAH, Complex Admixture History; MDE, Middle Eastern; iPD, idiopathic Parkinson's disease; HC, healthy controls; SD, standard deviation.

Table S2. Numbers and percentages of lyso-PRS SNPs identified in each ancestry.

| Ancestry | N | % |
| --- | --- | --- |
| European-GP2 | 101,090 | 94.0 |
| European-TUEPAC | 33,364 | 94.1 |
| AJ | 101,098 | 94.0 |
| AMR | 101,186 | 94.1 |
| SAS | 100,069 | 93.0 |
| CAS | 101,000 | 93.9 |
| AAC | 101,205 | 94.1 |
| AFR | 100,566 | 93.5 |
| EAS | 91,414 | 85.0 |
| CAH | 101,206 | 94.1 |
| MDE | 100,969 | 93.9 |

European-GP2, European-ancestry GP2 samples genotyped by NeuroBooster array; European-TUEPAC, European ancestry Tuebingen Parkinson cohort; AJ, Ashkenazi Jewish; AMR, Latino and indigenous Americas; SAS, South Asian; CAS, Central Asian; AAC, African American; AFR, African; EAS, East Asian; CAH, Complex Admixture History.

Table S3. lyso-PRS differences between iPD and HC across ancestries, adjusted by sex, age, and first five principal components.

| Ancestry | Controls | Patients | Beta | Standard error | P value |
| --- | --- | --- | --- | --- | --- |
| Idiopathic PD | | | | | |
| European-GP2 | 9,574 | 21,328 | 0.37 | 0.013 | 2.02e-172* |
| European-TUEPAC | 451 | 2,122 | 0.53 | 0.058 | 5.34e-20* |
| AJ | 220 | 1,169 | 0.31 | 0.074 | 2.33e-05* |
| AMR | 1,363 | 1,754 | 0.26 | 0.041 | 2.01e-10* |
| SAS | 304 | 212 | 0.26 | 0.089 | 2.93e-03* |
| CAS | 676 | 507 | 0.25 | 0.059 | 3.52e-05* |
| AAC | 595 | 266 | 0.083 | 0.082 | 0.31 |
| AFR | 2,580 | 1,145 | 0.066 | 0.037 | 0.075 |
| EAS | 1,448 | 2,486 | 0.14 | 0.037 | 2.47e-04* |
| CAH | 296 | 485 | 0.19 | 0.089 | 0.036* |
| MDE | 681 | 580 | 0.26 | 0.066 | 6.54e-05* |
| *GBA1*-PD (EUR) | | | | | |
| *GBA1*-PD vs iPD | 23,449 | 2,206 | 0.049 | 0.022 | 0.026* |
| *GBA1*-PD vs NMC | 482 | 2,206 | 0.45 | 0.051 | 2.05-18* |
| *GBA1*-PD vs HC | 10,025 | 2,206 | 0.44 | 0.025 | 7.83e-71* |

European-GP2, European-ancestry GP2 samples genotyped by NeuroBooster array; European-TUEPAC, European ancestry Tuebingen Parkinson cohort; AJ, Ashkenazi Jewish; AMR, Latino and indigenous Americas; SAS, South Asian; CAS, Central Asian; AAC, African American; AFR, African; EAS, East Asian; CAH, Complex Admixture History; MDE, Middle Eastern; HC, healthy controls; NMC, non-manifesting *GBA1* carriers.

Table S4. Summary statistics of lyso-PRS performance in the European population across *GBA1*-PD comparisons.

| **Groups** | **Threshold** | **PRS R2 adj** | **Full**  **R2** | **Null**  **R2** | **Coefficient** | **SE** | **No. of SNP** | **OR (95% CI)** | **AUC** | **Accuracy (95% CI)** | **Balanced accuracy** | **Sensitivity** | **Specificity** |
| --- | --- | --- | --- | --- | --- | --- | --- | --- | --- | --- | --- | --- | --- |
| *GBA1*-PD vs iPD | 0.1 | 0.0 | 0.018 | 0.017 | 0.049 | 0.022 | 4,060 | 1.05 (1.01 - 1.10) | 0.515 | 0.541 (0.535 - 0.547) | 0.513 | 0.481 | 0.546 |
| *GBA1*-PD vs NMC | 0.1 | 0.043 | 0.078 | 0.037 | 0.451 | 0.051 | 4,060 | 1.57 (1.42 - 1.74) | 0.621 | 0.548 (0.529 - 0.567) | 0.593 | 0.524 | 0.662 |
| *GBA1*-PD vs HC | 0.1 | 0.011 | 0.022 | 0.010 | 0.445 | 0.025 | 4,060 | 1.56 (1.49 - 1.64) | 0.621 | 0.577 (0.568 - 0.586) | 0.586 | 0.601 | 0.572 |

Column definitions were provided in Table 1. *GBA1*-PD, Parkinson’s disease with pathogenic *GBA1* variants; iPD, idiopathic Parkinson’s disease; NMC, non-manifesting *GBA1* carriers; HC, healthy controls.

Table S5. Results of random-effects meta-analysis for the lysosomal polygenic risk score (lyso-PRS) and clinical outcomes in European-GP2 cohort.

| Clinical outcomes | beta | se | 95%CI | P value | n_studies | I^2^ | CochransQ_pval | N_number |
| --- | --- | --- | --- | --- | --- | --- | --- | --- |
| Sex | 0.0094 | 0.016 | -0.022, 0.041 | 0.59 | 23 | 5.85% | 0.29 | 20,162 |
| Age at onset | -0.21 | 0.090 | -0.38, -0.03 | 0.022* | 18 | 5.36% | 0.16 | 17,733 |
| UPDRS-I | 0.48 | 0.18 | 0.13, 0.83 | 0.0073* | 3 | 0% | 0.42 | 1,100 |
| UPDRS-II | 0.87 | 0.23 | 0.42, 1.31 | 1.34e-04* | 3 | 0% | 0.84 | 1,105 |
| UPDRS-III | 1.13 | 0.50 | 0.15, 2.12 | 0.024* | 5 | 50.46% | 0.045 | 1,847 |
| UPDRS-IV | 0.32 | 0.12 | 0.082, 0.57 | 0.0088* | 3 | 0% | 0.25 | 736 |
| Cognition | 0.082 | 0.047 | -0.011, 0.17 | 0.083 | 6 | 0% | 0.60 | 3,241 |
| BDI-II | -0.0075 | 0.21 | -0.41, 0.40 | 0.97 | 3 | 0% | 0.62 | 1,398 |
| GDS-15 | 0.19 | 0.15 | -0.10, 0.48 | 0.21 | 2 | 54.83% | 0.14 | 871 |
| RBDSQ | 0.18 | 0.11 | -0.030, 0.39 | 0.093 | 2 | 0% | 0.68 | 871 |
| ESS | 0.16 | 0.10 | -0.035, 0.36 | 0.11 | 4 | 8.81% | 0.42 | 2,109 |

UPDRS, Unified Parkinson's Disease Rating Scale; BDI-II, Beck Depression Inventory II; GDS-15, Geriatric Depression Scale-15; RBDSQ, REM Sleep Behaviour Disorder Screening Questionnaire; ESS, Excessive Daytime Sleepiness.

Table S6. Results of regression analysis between the lysosomal polygenic risk score (lyso-PRS) and clinical outcomes in European-TUEPAC cohort.

| Clinical outcomes | beta | se | p value | N_number |
| --- | --- | --- | --- | --- |
| Sex | 0.064 | 0.043 | 0.14 | 2,122 |
| Age at onset | -0.38 | 0.26 | 0.14 | 2,075 |
| UPDRS-I | -0.30 | 0.25 | 0.23 | 591 |
| UPDRS-II | -0.25 | 0.34 | 0.47 | 579 |
| UPDRS-III | 0.25 | 0.36 | 0.45 | 1,670 |
| UPDRS-IV | 0.22 | 0.11 | 0.045* | 1,301 |
| Cognition | -0.17 | 0.058 | 3.58e-03* | 1,646 |
| BDI-II | -0.10 | 0.22 | 0.63 | 1,512 |

UPDRS, Unified Parkinson's Disease Rating Scale; BDI-II, Beck Depression Inventory II.

Table S7. Results of random-effects meta-analysis for the lysosomal polygenic risk score (lyso-PRS) and clinical outcomes in European-GP2 cohort after adjustment for disease duration.

| Clinical outcomes | beta | se | 95%CI | P value | n_studies | I^2^ | CochransQ_pval | N_number |
| --- | --- | --- | --- | --- | --- | --- | --- | --- |
| UPDRS-I | 0.10 | 0.23 | -0.36, 0.56 | 0.67 | 2 | 0 | 0.92 | 573 |
| UPDRS-II | 0.64 | 0.27 | 0.11, 1.17 | 0.018* | 2 | 0 | 0.85 | 575 |
| UPDRS-III | 0.40 | 0.59 | -0.75, 1.55 | 0.50 | 3 | 55.93% | 0.099 | 911 |
| UPDRS-IV | 0.36 | 0.13 | 0.10, 0.63 | 6.20e-03* | 2 | 0 | 0.38 | 450 |
| Cognition | 0.037 | 0.058 | -0.077, 0.15 | 0.52 | 5 | 0 | 0.66 | 2,541 |
| BDI-II | 0.15 | 0.28 | -0.40, 0.69 | 0.60 | 2 | 0% | 0.53 | 924 |
| GDS-15 | -0.026 | 0.12 | -0.26, 0.21 | 0.83 | 1 | / | / | 366 |
| RBD | -0.15 | 0.17 | -0.49, 0.18 | 0.38 | 1 | / | / | 366 |
| ESS | 0.050 | 0.22 | -0.38, 0.47 | 0.82 | 2 | 33.99% | 0.22 | 682 |

UPDRS, Unified Parkinson's Disease Rating Scale; BDI-II, Beck Depression Inventory II; GDS-15, Geriatric Depression Scale-15; RBDSQ, REM Sleep Behaviour Disorder Screening Questionnaire; ESS, Excessive Daytime Sleepiness.

Table S8. Results of regression analysis between the lysosomal polygenic risk score (lyso-PRS) and clinical outcomes in European-TUEPAC cohort.

| Clinical outcomes | beta | se | p value | N_number |
| --- | --- | --- | --- | --- |
| UPDRS-I | -0.30 | 0.25 | 0.23 | 591 |
| UPDRS-II | -0.23 | 0.31 | 0.46 | 579 |
| UPDRS-III | 0.11 | 0.32 | 0.74 | 1,670 |
| UPDRS-IV | 0.18 | 0.098 | 0.070 | 1,301 |
| Cognition | -0.16 | 0.058 | 4.57e-03* | 1,646 |
| Depression | -0.12 | 0.22 | 0.58 | 1,512 |

UPDRS, Unified Parkinson's Disease Rating Scale; BDI-II, Beck Depression Inventory II.

Figure S1. Forest plots display regression estimates with 95% confidence intervals for the associations between lyso-PRS and non-motor scales across the European-GP2 training cohort.

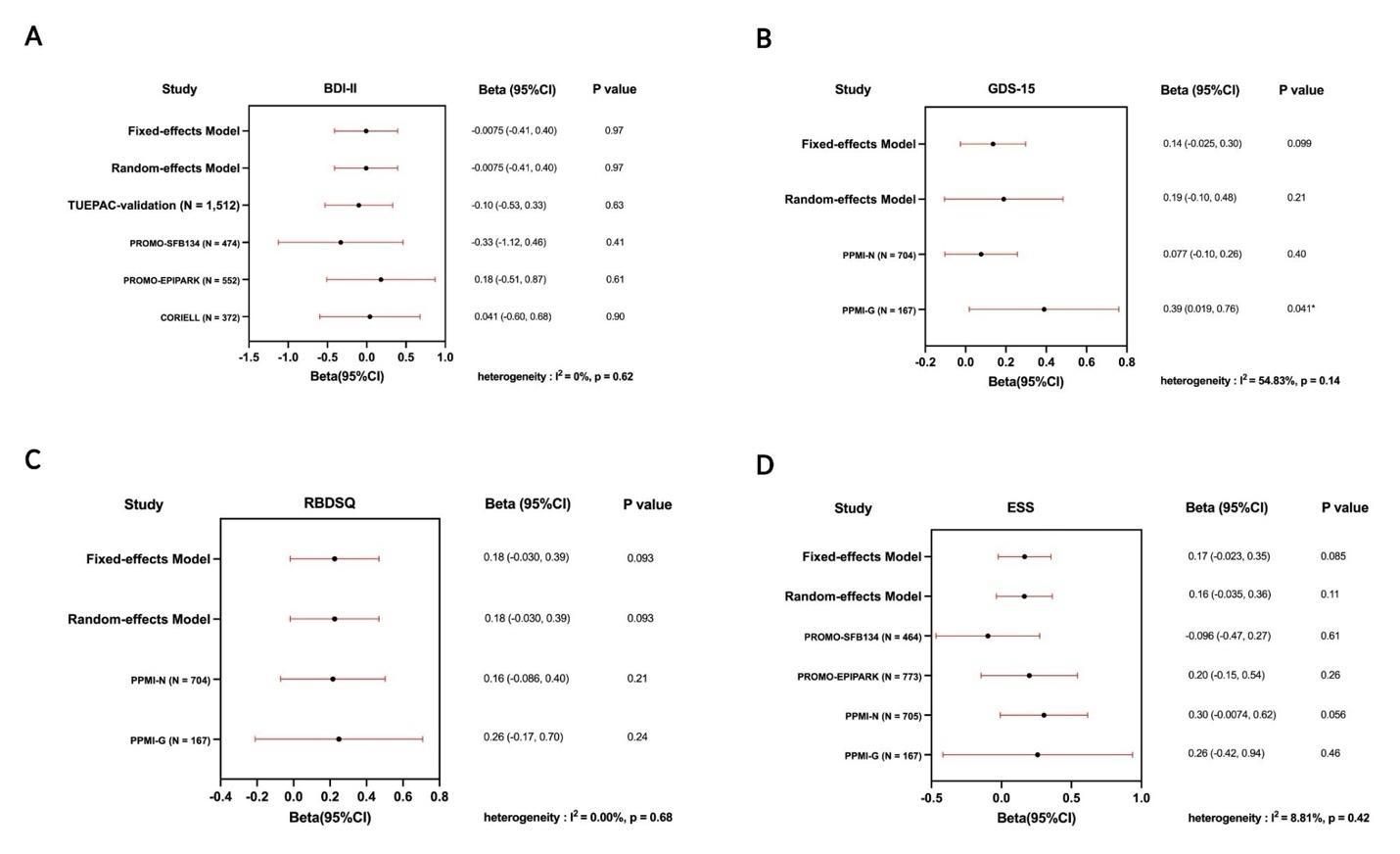

(A) Depression; (B) GDS-15; (C) RBDSQ; (D) ESS. GDS-15, Geriatric Depression Scale; RBDSQ, REM Sleep Behavior Disorder Screening Questionnaire; ESS, Epworth Sleepiness Scale.

Figure S2. Forest plots summarize regression estimates with 95% confidence intervals for the association between lyso-PRS and clinical outcomes across European-GP2 training cohort and European-TUEPAC validation cohort after adjustment for disease duration.

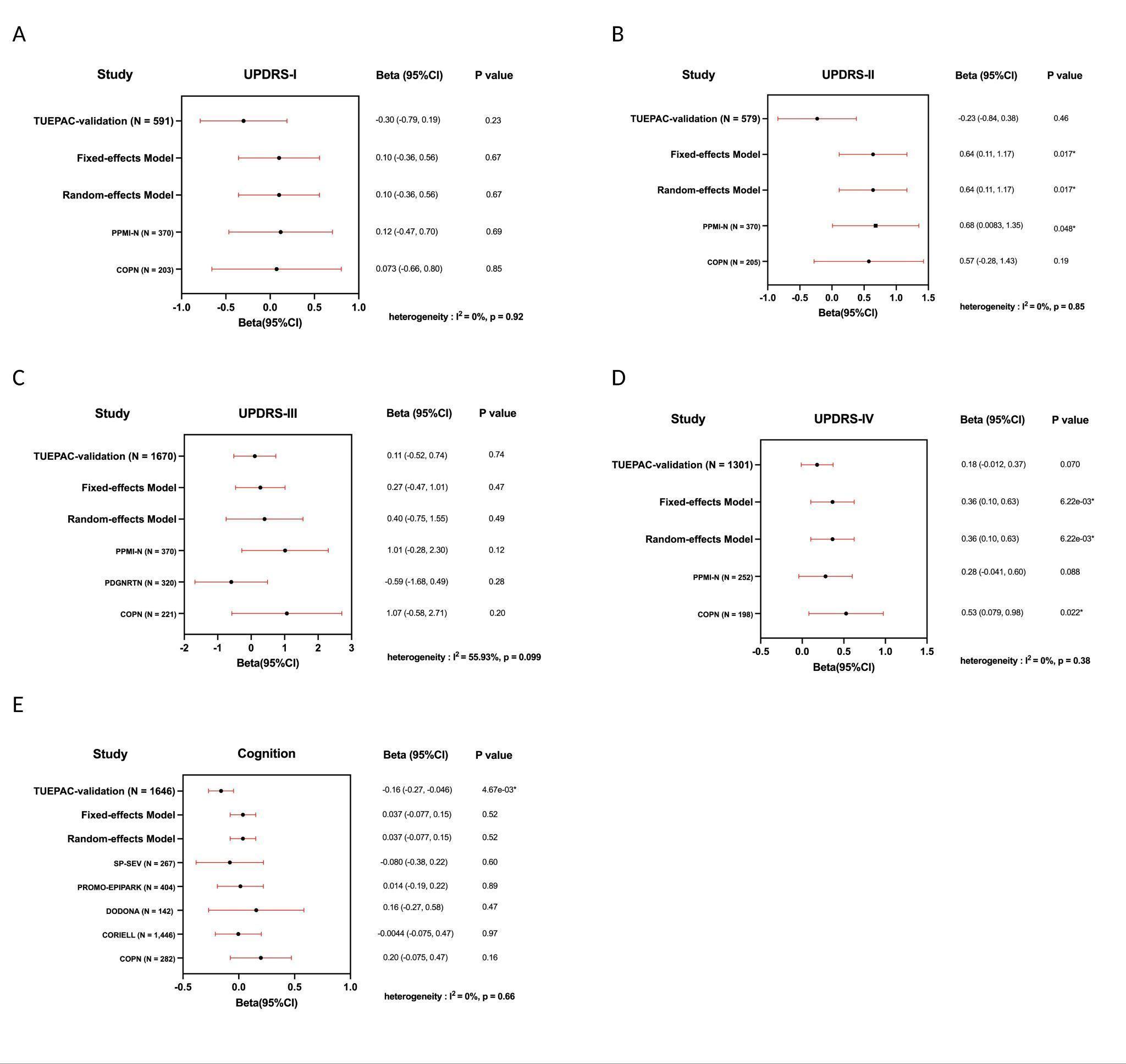

(A-D) UPDRS I-IV; (E) Cognition. Regarding cognitive impairment analyses, PD patients were classified as normal cognition or impaired cognition based on established cutoffs for the Mini-Mental State Examination (MMSE, <24), Montreal Cognitive Assessment (MoCA, <26), and Scales for Outcomes in Parkinson’s Disease-Cognition (SCOPA-COG, <24). European-GP2, GP2 samples in the European population genotyped using the Illumina NeuroBooster Array; European-TUEPAC, European ancestry Tuebingen Parkinson cohort; UPDRS, Unified Parkinson's Disease Rating Scale.
